# Improve data management in register-based research: Transition from CSV to Parquet

**DOI:** 10.1101/2025.10.15.25337992

**Authors:** Simone Rahel Fenk, Kari Furu, Inger Johanne Bakken

## Abstract

**Aims:** To identify an efficient file format for data delivery from large administrative registers for research, testing the full workflow from data extraction to research data management.

**Methods:** Through collaboration between a data delivery and a research department within the same institute, we evaluated each step from data extraction and delivery to management and usage, comparing CSV and Parquet formats.

**Results:** Switching from the unstructured, text-based CSV format to the highly structured Parquet format significantly optimized all processes by reducing file sizes and saving processing time. The Parquet format also provided access to advanced data management techniques, simplifying further work. Despite these advantages, the basic programming required for Parquet format is not very different from that for CSV. We provide a tutorial and examples as online supplement.

**Conclusions:** We strongly recommend replacing CSV files with contemporary data formats. The Parquet file format proved to be an excellent option throughout the entire process from data extraction to research implementation

## Introduction

The Nordic countries are known for their population-based health registers. By means of the personal identity number (PIN) unique to every citizen, registers can be linked to each other and to other data sources, such as census data and information on education, income, and family structure.^1^

The medical birth registers and the cause-of-death registers contain information on one-time events in a person’s life. Other registers, like the medical quality registers, are usually dedicated to conditions or treatments that are relatively rare. Even though such registers are population-based, size is not usually an issue in research when it comes to data management or analysis.

The administrative health registers, however, contain broad information on all health care utilization in the country and rapidly accumulate large amount of data over time. For example, the Norwegian Patient Registry (NPR) and the Norwegian Registry of Primary Health Care (NRPHC) contain information on all inpatient and outpatient encounters in specialist health care from 2008 and in primary health care from 2016, respectively.^2^ The Norwegian Prescribed Registry (NorPD) holds data on all prescription fills at Norwegian pharmacies from 2004 onwards.^3^ For context, during a typical month in 2024, there were approximately 1.4 million general practitioner’s consultations (recorded in NRPHC) and 0.8 million outpatient visits at somatic hospitals (recorded in NPR).^4, 5^ These data represent only a small fraction of what is reported to NRPHC and NPR. Although NorPD does not present online statistics on the total annual number of prescriptions filled, it reports that more than 4 million people (75.3 % of the population) filled at least one prescription in 2024, with defined daily doses surpassing three billion.^6^

Data in modern, well-functioning registers are highly organized and well-structured, with substantial human expertise and advanced technological resources dedicated to maintaining up-to-date scalable registers. The possibilities and advantages of using data from the mandatory, population-based health registers in the Nordic countries for research purposes are well-documented.^7, 8^ Studies often apply advanced statistical methods within carefully planned study designs, such as the increasingly popular target trial emulation approach.^9^

These advanced and modern efforts differ notably from the workflow when data are extracted from databases and delivered to researchers. In our experience, the data format for transfer from registry to research project has not been changed since the early days of research using data from administrative registries.

In this paper, we demonstrate how transitioning from a traditional unstructured row-based data file format to a contemporary structured column-oriented data file format significantly enhances data storage efficiency and improves data management capabilities. Our case is a large research project on pregnancy and medication use, Drugs in Childhood and Reproductive Life with second author (KF) as a project leader.^10,11^

## Materials and Methods

In this collaboration between a data delivery department and a research department, the first author (SRF) was responsible for data delivery from the databases (NPR and NRPHC), while the last author (IJB) had data management and research responsibilities. This setting is important as we covered the entire workflow from data extraction and data delivery to data management and use of data in research.

By request at the data delivery department, research data is extracted from databases according to the study population and variable list specified by the researchers, ensuring that data is tailored to the project’s unique requirements (see Figure 1). The final data is extracted and stored in a suitable format, traditionally usually the text-based, unstructured CSV format. Since the data files included sensitive data and consisted of a large data set, data files need to be compressed and encrypted before delivery via a file sharing system.

Drugs in Childhood and Reproductive Life holds approvals for updates until 2032. The purpose of the research project is to study drug use in general, and short-term and long-term safety related to drug use in the population, but with a special focus on safety during pregnancy and in children/adolescents. The study population is defined from a combination of the Norwegian Population Register and the Norwegian Medical Birth Registry (MBRN): all women and men registered in the Norwegian Population Register as born from 1944 onwards, supplemented with individuals only registered in the MBRN. The total population consists of more than 9 million individuals. Data for this population are extracted and delivered to the project from numerous data sources, including NRPHC, NPR, and NorPD.

In winter of 2024, data from NPR, covering the period from 2008 to August 2024, and data from the NRPHC spanning from 2016 to August 2024, were delivered in the traditional row-based format CSV (Delivery 1).

We began exploring alternatives to CSV for delivering large datasets and quickly selected the Parquet file format for further work. We identified numerous online resources, referencing only a few that we found most beneficial.^12-15^ Importantly, we also engaged with scientists at other institutions experienced in handling large datasets to discuss whether Parquet could effectively address our CSV challenges (see acknowledgements).

In spring 2025, a new data delivery was completed using the Parquet file format, which included updated data covering the entire year 2024 (Delivery 2). We will discuss the differences between CSV and Parquet and highlight the tools we have found most useful.

This methodology improvement project was carried out in accordance with regulations of the NRPHC and NPR and did not require ethical approvement. The Drugs in Childhood and Reproductive Life Project was approved by the Committee for Medical and Health Research Ethics in Norway (REK South- East A; 2017/2546) and The Norwegian Data Inspectorate in Norway (17/02068/Norwegian Data Inspectorate).

## Results

The first data delivery from NPR and NRPHC was provided in the traditional CSV format with data updated until August 2024, while the second delivery was in Parquet format with data updated until December 2024 (Table 1).

**Table.** Comparison of data deliveries 1 and 2 in the Drugs in Childhood and Reproductive Life project.

|  | Data delivery 1 | Data delivery 2 |
| --- | --- | --- |
| **Time of delivery** | Autumn 2024 | Spring 2025 |
| **Content** | NPR 2008 – August 2024 | NPR 2008 – 2024 |
|  | NRPHC 2016 – August 2024 | NRPHC 2016 – 2024 |
| **Format** | CSV | Parquet |
| **Size** | 76.24 GB | 12.97 GB |
| **Partitioning** | Produced yearly files through standard loop programming | Used in-built options for partitioning by year for creating a Parquet file system. |
| **Overall structure** | CSV files split by year, each requiring individual handling. | A Parquet file system that functions like a unified single file. |
| **Storage** | Row-based | Hybrid-based (rows grouped by row groups, then column partitioned) |
| **Metadata** | None, all data stored as text. | Metadata is stored within the file, preserving the original database format |
| **Compression** | Time-consuming and resource-intensive process using 7-zip | In-built compression |
| **Password encrypting** | 7-zip | 7-zip |
| **Processing\* method** | Reading and processing individual files (data.table package) | Querying a Parquet file system (arrow package) |
| **Processing time** | 27 minutes | 28 seconds |

NPR: Norwegian Patient Registry; NRPHC: Norwegian Registry for Primary Health Care
\* Calculating the number of individuals in Norway with a prescription from 2004 to 2024 using data from the Norwegian Prescribed Drug Registry.

Despite the increased information **content** in Delivery 2, with Parquet file **format** rather than CSV, the total file **size** for NPR and NRPHC data combined was reduced from 76.24 GB to 12.97 GB (-83%).

Using looping techniques for splitting up data into smaller files became unnecessary when using Parquet, as Parquet data easily can be organized as Parquet file systems by taking advantage of the **partitioning** options. In our example, we partitioned data by year, but it is also possible to use multiple categorical variables for partitioning. Thus, while the **overall structure** for Delivery 1 was a set of CSV files, Delivery 2 consisted of a Parquet file system. Parquet file systems comprise multiple files that are interconnected through their intrinsic structure, offering an experience as if working with a single cohesive file. Partitioning not only simplifies data management but also accelerates data processing. For instance, a query targeting a single year will only access that specific part of the system, making the querying process highly efficient.

In addition to partitioning, the columnar **storage** format in Parquet also allows for faster data retrieval and processing, meaning that in-memory tasks can be tailored to only using the data necessary, within the structure provided directly from the register owners.

Since CSV files are text-based, they lack intrinsic **metadata** regarding data types. In contrast, Parquet files include additional metadata specifying the format of each column (string, date, integer, numeric), along with statistics for each column and further information about offset and size.^16^ In our example, when extracting data from register databases for research purposes, the original data formats are preserved in the resulting Parquet file system. This allowed us to leverage the careful curation of the original databases.

While the writing time to Parquet was comparable to CSV, Parquet’s intrinsic and advanced **compression** provided significant savings in time and resources on the register delivery side. Parquet files where password **encrypted** by using the standard 7-Zip program in the same way as the CSV files.^17^

While data from NPR and NPRHC were delivered as Parquet files in Delivery 2, only CSV files were delivered from NorPD, representing all prescription fills in Norway from 2004 to 2024, split into yearly files. We converted these CSV files into a Parquet file system, partitioned by year, mirroring the structure used in the NPR and NPRHC deliveries. In this process, we used the DuckDB package in R and manually specified the data format for each relevant column. To minimize the programming workload, we included only a few key variables essential to the research project. This effort was rewarding, as it not only reduced the total file size from 146 GB to 6 GB but also provided access to powerful data handling tools not available for CSV files, optimizing our ability to efficiently query this large dataset.

Since NorPD data were originally delivered as CSV files, we had the opportunity to compare processing time using a straightforward **example:** calculating the number of individuals in Norway with a prescription fill from 2004 to 2024 with two different **methods**. With the original CSV files, we read each file into memory using *fread* from data.table, extracted distinct personal identifiers, and then aggregated them to determine the total number of individuals over the entire period. In contrast, querying the Parquet file system using the arrow package required no reading into memory and allowing for a comprehensive query in a single operation. The difference in **processing time** was striking: 27 minutes using the CSV files and 28 seconds with the Parquet file system.

Both methods yielded the same result. We provide the code for the processing time test in the Online Supplement.

## Discussion

We found the advantages of using the Parquet file format evident from the outset. We quickly integrated Parquet into our daily routine. Furthermore, we implemented these changes across our respective departments within just a few months.

### Strengths and limitations of the present study

The primary strength of this study is our extensive experience with data handling and management, both on the register holder side and on the research side. We successfully tested the transfer and utilization of research data from NPR and NRPHC as Parquet files and converted large CSV data to Parquet. The Parquet data structures from NPR and NRPHC served as excellent templates for converting the NorPD data, which was delivered as CSV.

We have only worked with SQL and R, which represents a significant limitation of the current study.

### Implementation

Parquet is an open-source data file format, making it accessible for everyone. It has become a de facto standard for storage and sharing large tabular data sets across a variety of systems and analytic tools.^18^ However, we recognize that Parquet may present challenges in traditional register-based research. While Parquet file handling is supported in R and Python, using Parquet files in other statistical software like SPSS or Stata might not be straightforward, potentially presenting difficulties for researchers who prefer these programs.

Beyond the potential challenges of adapting to new data handling tools, research projects, such as our case study, often require repeated data updates. Some researchers may argue that receiving data in a different format could disrupt established workflows and add complexity. However, in our view, the advantages of using Parquet greatly outweigh the initial extra effort required. In the online supplement, we provide examples of initial steps for handling Parquet file systems using the R package arrow. These methods are accessible and similar to those found in the widely used R package tidyverse.

## Conclusions

We found that switching from the row-based, unstructured CSV format to the highly structured, columnar Parquet format significantly improved efficiency in terms of file size and processing time both at the data delivery side and at the research department site. Utilizing Parquet and associated R packages swiftly became an integral part of our routine workflow, and we recommend those working with large datasets in CSV to consider transitioning to Parquet, preferably in a program that will take the full advantage of the possibilities of this data structure. Another possibility is to learn the basic data management steps of Parquet data handling in R or Python, and then to continue the usual workflow.

## Data Availability

Data are accessible to authorized researchers after ethical approval and application to https://helsedata.no/. We also used simulated data that can be reproduced as described in the supplemental material.

## Acknowledgements

We wish to thank IT-consultant Francesco Frassinelli and Dr. Sara Ghaderi at the Norwegian Tax Administration for discussions, insightful advice and encouragement in the initial phases of this work.

## Online Supplement 1

### Tutorial: Arrow and Parquet

**Objective:** To lower the entry barrier for those interested in working with Parquet files. We use a fake data set.

### Introduction

Please check that packages `arrow`, `data.table` and `dplyr` have been installed.

The first code block activates the necessary packages and generates a random data set with 1 million rows.

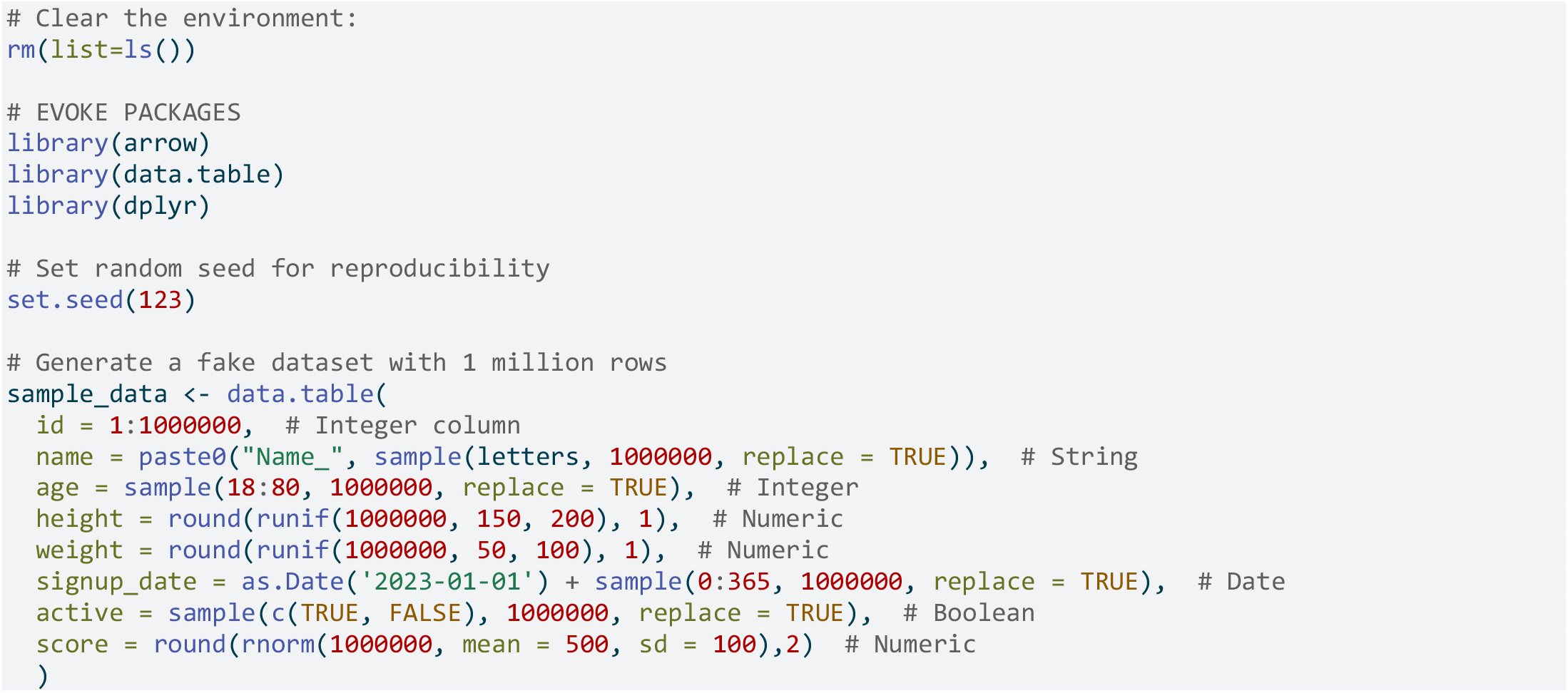

We now designate file paths for a CSV file and a Parquet file and write the files to disk.

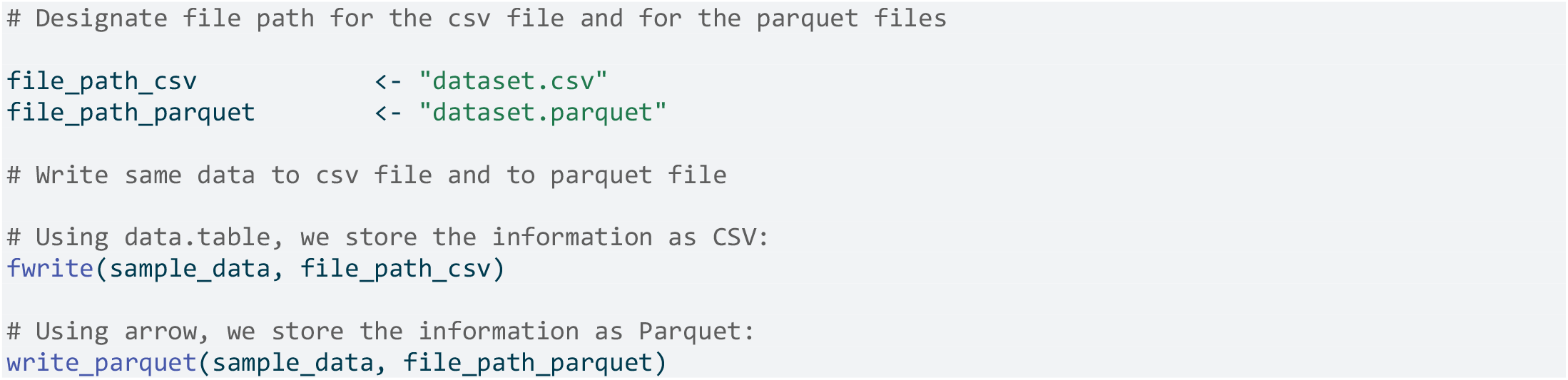

After this step, you should have a new file in your working directory, “dataset.csv” and a new folder “dataset.parquet”.

We can compare file sizes by inspecting the folder by using the `file.size` command.

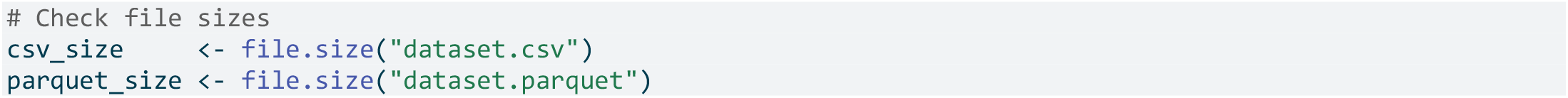

Note that the Parquet file size is just 22.7 percent of the CSV file size.

We clean up the environment again:

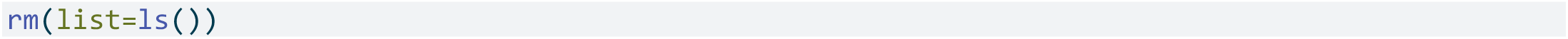

We now only use the Parquet file in the further work and start taking advantage of the `arrow` package.

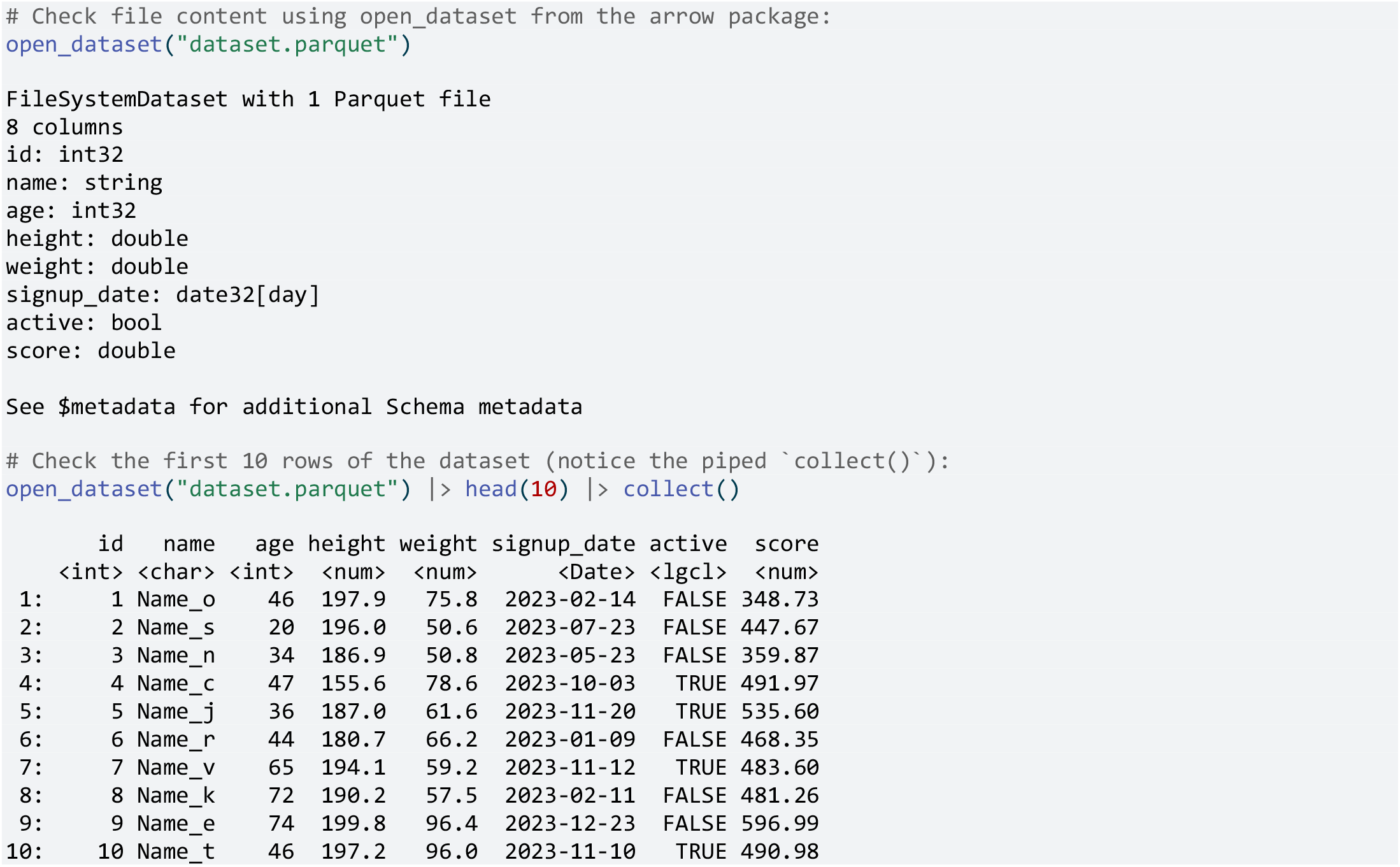

Now repeat the above command, piping in a select (notice logic of the code and the similarity to `tidyverse`).

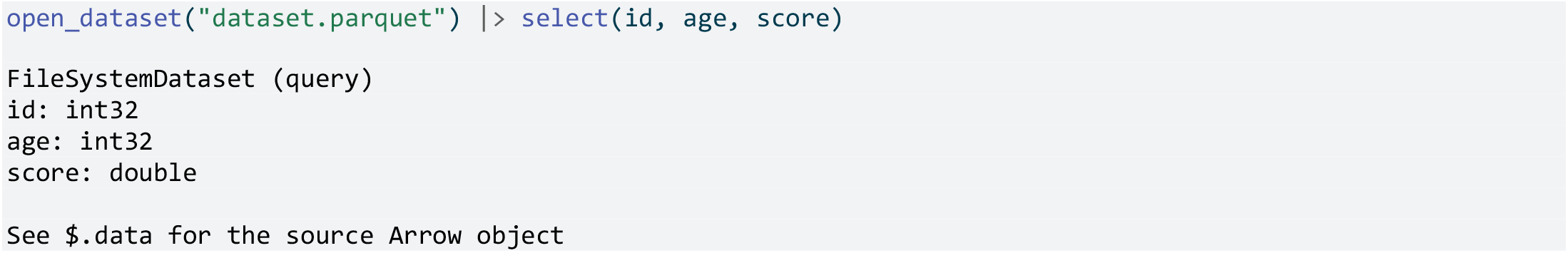

Now we want to add a filter as well. Again, use pipe operations and inspect the result in the console and the environment:

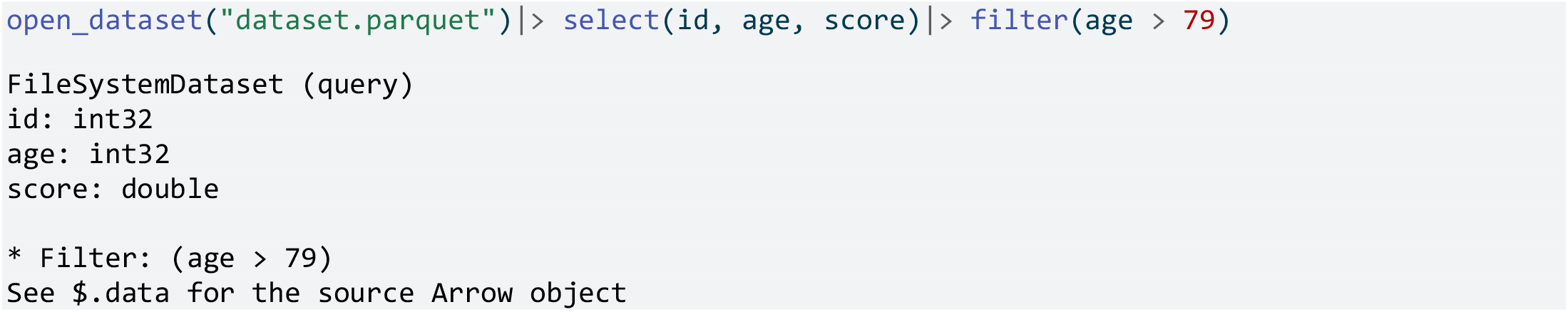

We want part of our data into the environment: The variables `id`, `age` and `score` for people above 80 years. Notice `collect()` as the last piped command:

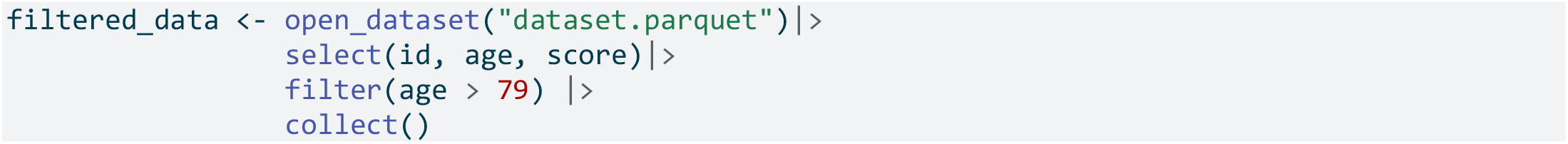

Now in the environment is a small fraction of the “original” data, selected three variables and filtered to a part of the population.

Notice that the full data set never was read into the environment.

Using `data.table` the same operation would have been:

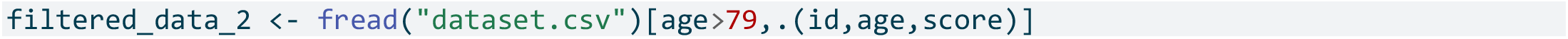

Or by in `dplyr` :

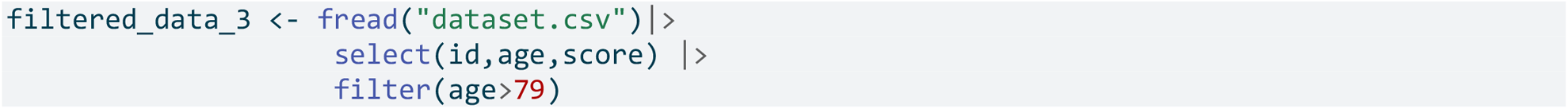

It is time for cleaning up the environment again:

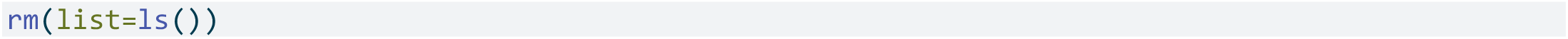

### Partitioning and compression

Previously, we saved our fake data set to a Parquet file without partitioning.

Now we aim to save data to a **Parquet file system**, partitioned by a categorical variable. Let’s begin by creating a simple fake data set, this time consisting of 10 million rows.

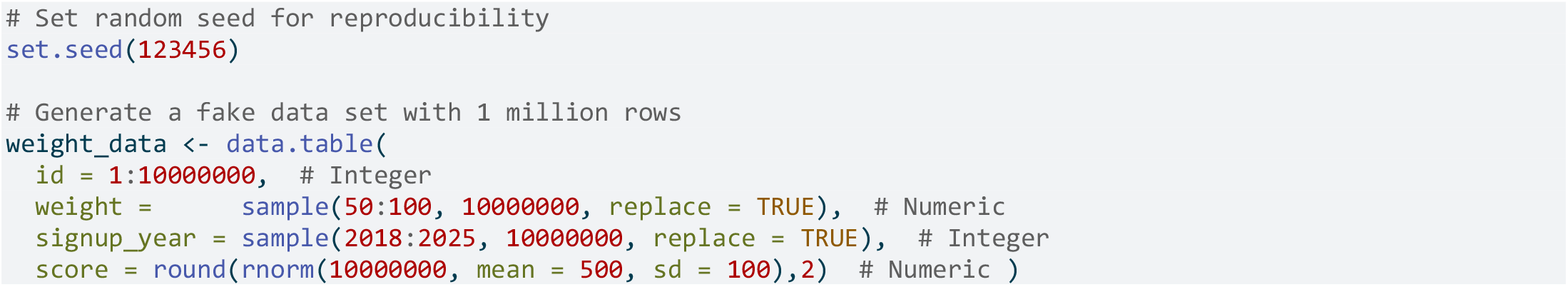

We will save this data set as a Parquet file system, using Zstandard (ztsd) compression and partitioning by the variable for year of signup.

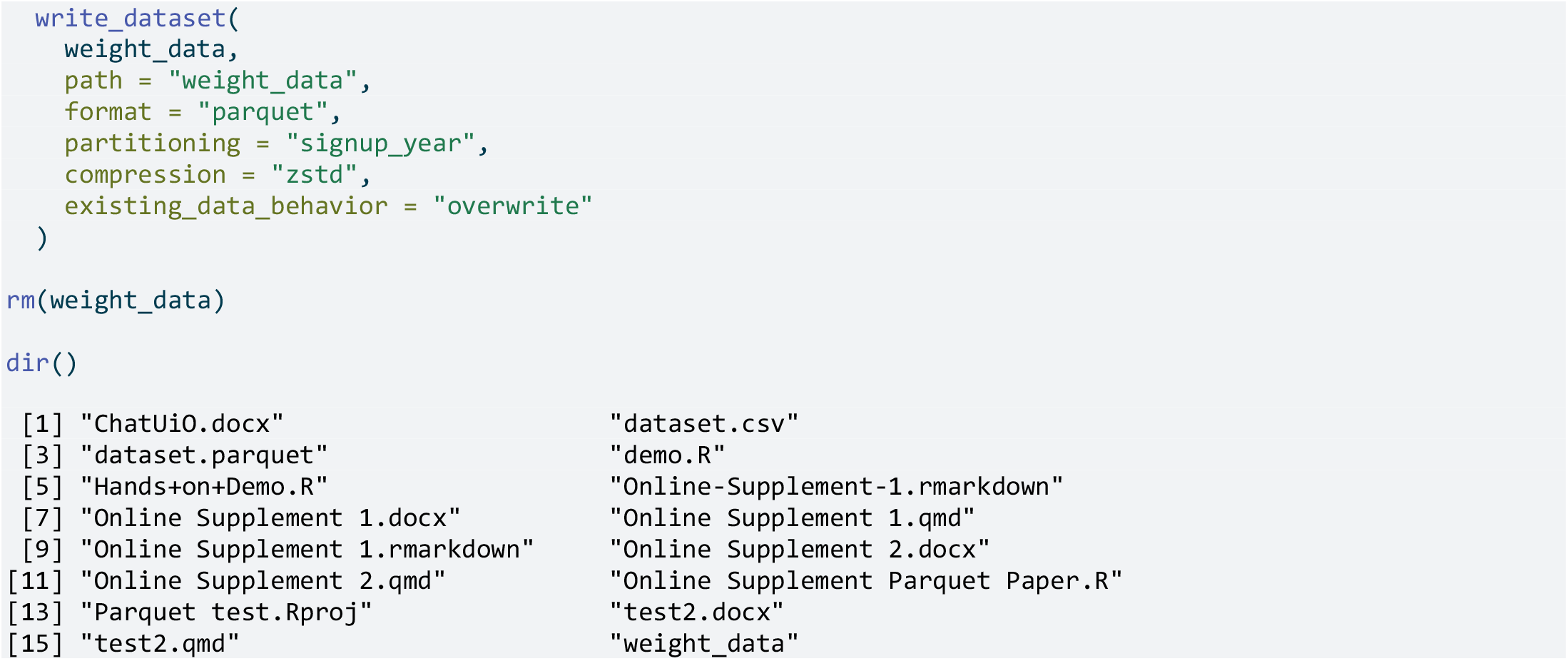

We see that there is now a folder called “our_parquet_system” in the the directory.

Let’s take a look at the content of the folder:

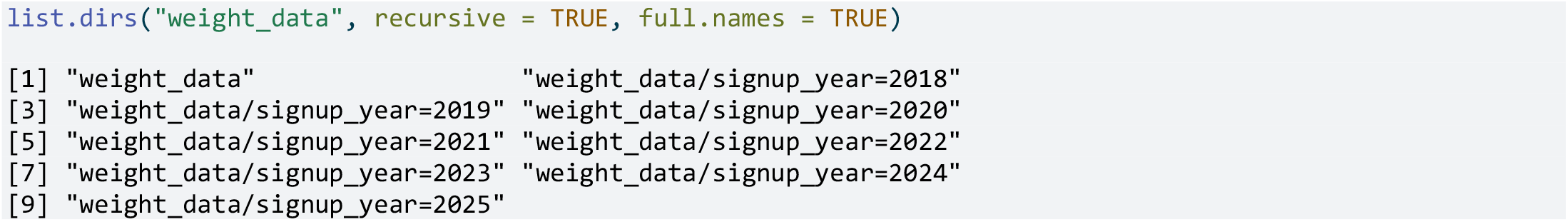

We observe that there are eight subfolders, one for each sign-up year. We will now take advantage of the combined capabilities of `arrow`and Parquet in a few simple examples.

First, check the overall content:

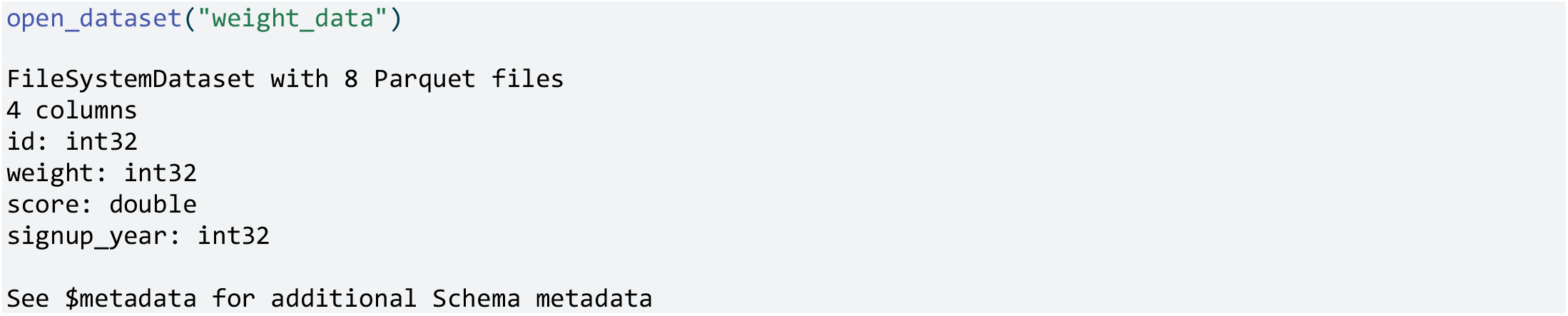

We learn about the variables and the overall structure from the above command (without reading the file into memory).

Now, we aim to conduct our first analyses, focusing on gathering the following information:

- The total number of distinct ID’s (although we already know the answer)
- The average weight by year
- The total number of distinct ID’s just for 2025

We gather this information as objects in the environment.

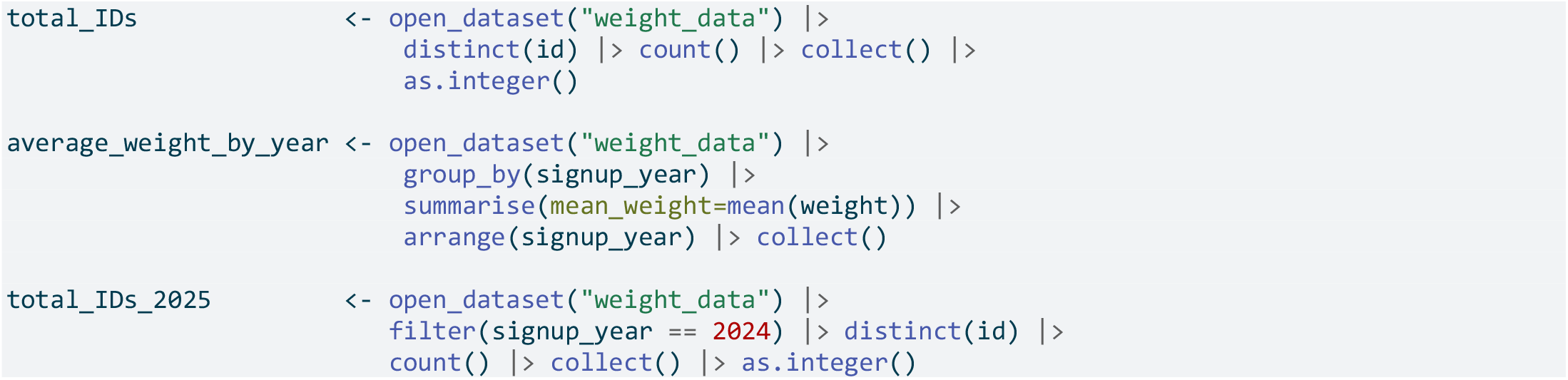

We learnt that there were 10 000 000 people in the sample in total. In 2025, a total of 1 249 854 signed up.

The weight was no surprise extremely stable over the years:

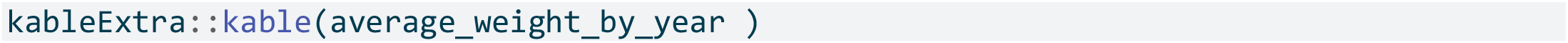

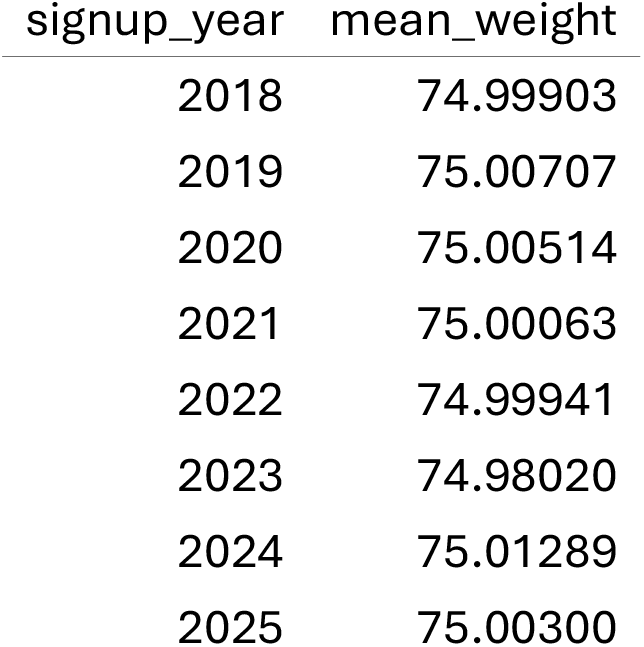

| signup_year | mean_weight |
| --- | --- |
| 2018 | 74.99903 |
| 2019 | 75.00707 |
| 2020 | 75.00514 |
| 2021 | 75.00063 |
| 2022 | 74.99941 |
| 2023 | 74.98020 |
| 2024 | 75.01289 |
| 2025 | 75.00300 |

## Conclusions

We hope that this tutorial can help you get started with arrow and Parquet.

## Online Supplement 2

### Source Code for our Processing Time Example

**Example:** We checked processing time for calculating the total number of people in Norway with a prescription in Norway using 1) Arrow on a Parquet file system created from the delivered CSV files 2) data.table on CSV files delivered from the Norwegian Prescription Database.

Run time was 28 minutes for the first approach and 27 seconds for the second approach. Please note that this supplement only gives the source code (cannot be run)

### Source Code for Speed Test

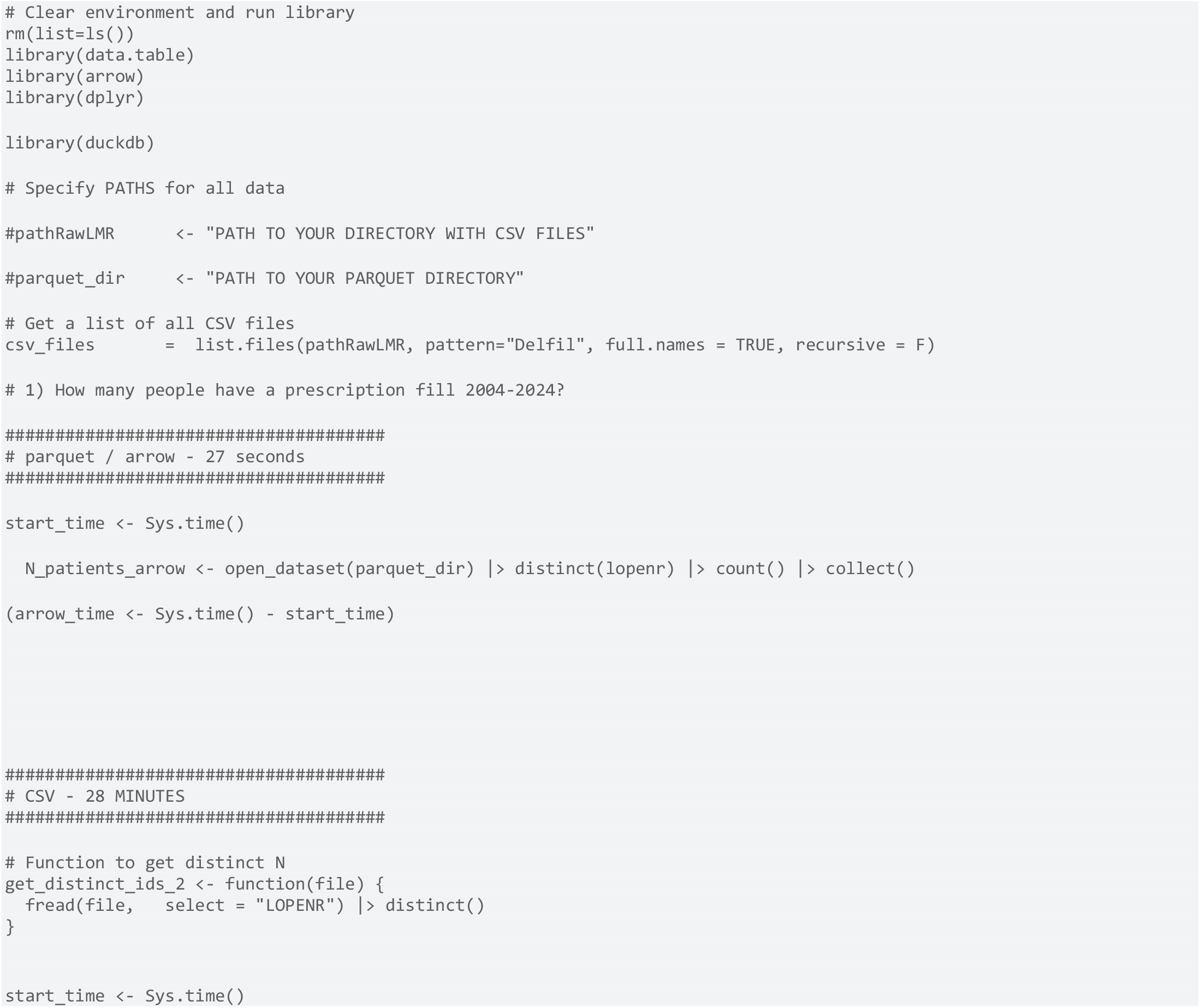

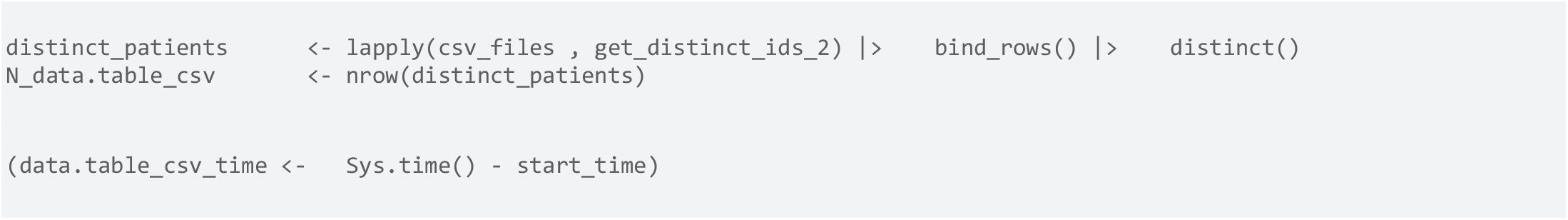

### Translating Data from CSV to Parquet using duckdb

DuckDB is a bit more complicated to work with than arrow, but comes in handy when working with particularly large data sets. Please note that this part only gives example code and can’t be run.

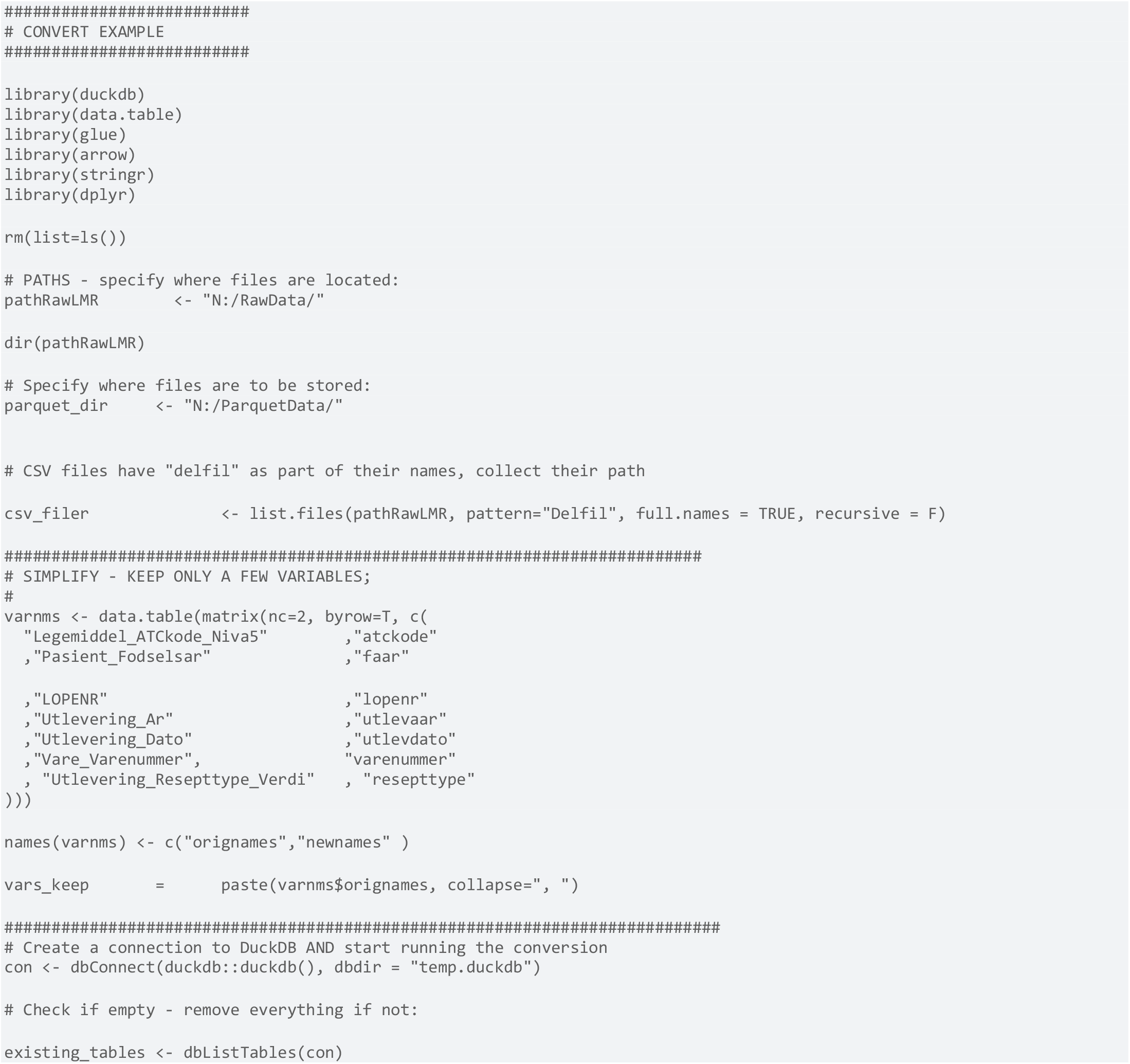

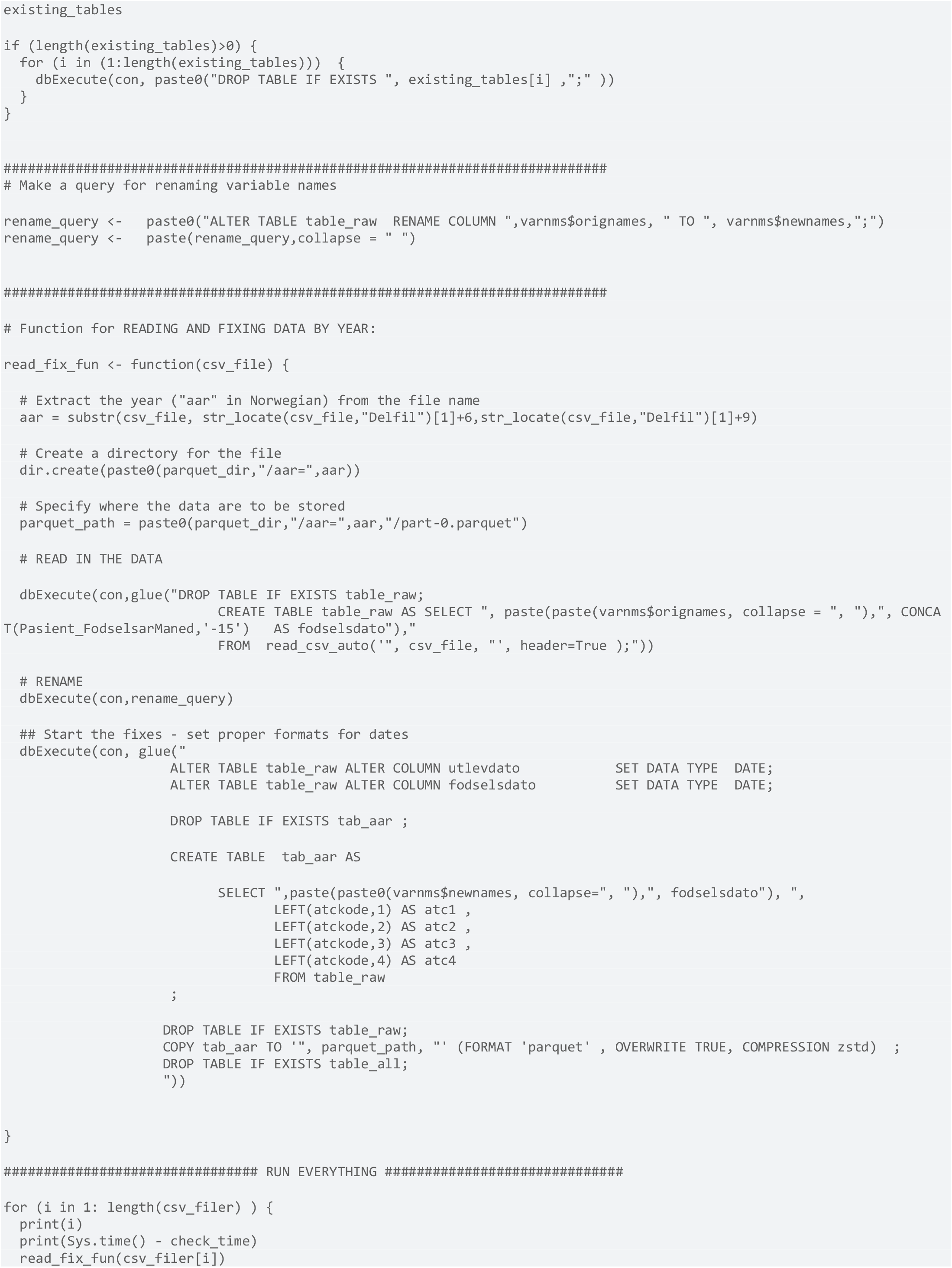

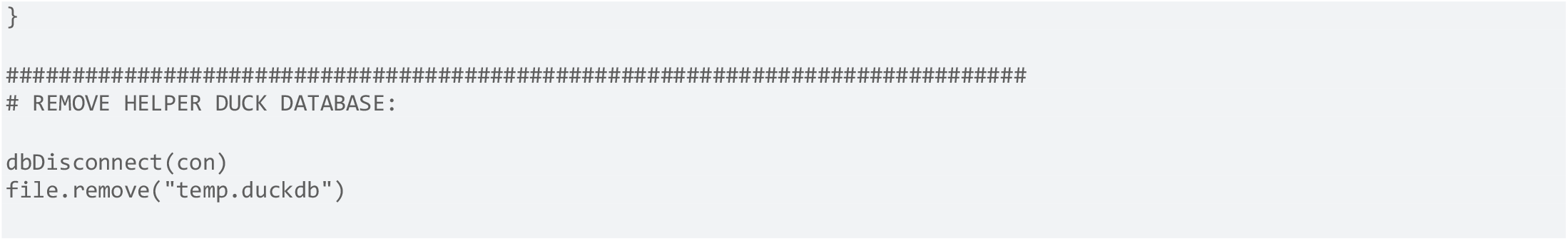

